# Schistosomiasis prevalence associated with water catchment geography in a rice farming community in South-eastern Madagascar: A cross-sectional exploratory study

**DOI:** 10.64898/2026.08.25.26361342

**Authors:** James Bevan, Nina Finley, Herizo Randrianandrasana, Tanguy Felistino Razafiarimihoby, Andry Tsirimanana, Elinambinina Rajaonarifara, Ryan Parks, Lena Reiter, Tahinamandranto Rasamoelina, Sakib Burza, Laura Braun

## Abstract

**Background:** Madagascar has one of the highest burdens of schistosomiasis globally, yet epidemiological evidence from the South-east of the country remains limited. Existing surveillance estimates suggest relatively low prevalence in this region. This contrasts with the region’s environmental suitability for transmission and its population’s high occupational risk through rice farming.

**Methods:** A cross-sectional exploratory study was conducted across 13 villages around the Manombo Special Reserve, Farafangana District, between May and July 2025. Participants were surveyed for geographic, environmental and behavioural risk factors for schistosomiasis infection. Infection was diagnosed by Kato-Katz stool microscopy. Multivariable mixed-effects logistic and linear regression, with village as a random intercept, estimated associations with infection and intensity respectively, adjusting for the water catchment area of household water sources, sex, age group, recent praziquantel treatment and village-level open defecation rate.

**Results:** 227 participants were included. Overall *S. mansoni* prevalence was 53.3% (95% CI 46.8–59.8%); median infection intensity was 96 (IQR 48–288) eggs per gram of stool among those infected. Surface water contact was near-universal (97.3%). Prevalence varied markedly between neighbouring villages. No demographic, behavioural, or WASH variable was independently associated with infection. The water catchment area of household water sources was the primary predictor of infection.

**Conclusions:** Prevalence substantially exceeded current surveillance estimates, suggesting this population is underserved by existing control programmes. Water catchment area was the dominant determinant of transmission heterogeneity, possibly consistent with *Biomphalaria* habitat suitability. Water catchment area may represent a valuable and underutilised unit for targeting schistosomiasis surveillance and control in Madagascar.

## Background

Schistosomiasis, a parasitic helminth infection, is estimated to affect approximately 150 million people worldwide.^1^ Contracted through contact with infested water, it is a disease of poverty associated with poor access to water, sanitation and hygiene (WASH) services and accounts for approximately 1.6 million disability-adjusted life years (DALYs) which occur almost entirely in low-and-middle income countries (LMICs), particularly in sub-Saharan Africa.^2^ Madagascar exemplifies the intersection of this burden with poverty, globally it has the fifth highest estimated schistosomiasis prevalence, while simultaneously experiencing the fifth lowest by GDP per capita.^3,4^ The disease exacerbates these inequities ranking second among parasitic infections globally in terms of socioeconomic impact.^5^

In Madagascar, *Schistosoma mansoni* (*S. mansoni*) predominates in the south and east of the country, and *S. haematobium* in the north and west of the country, with some areas of co-endemicity.^6^ This pattern is thought to be largely driven by their different respective intermediate snail hosts which require different environmental conditions for survival and reproduction, including water body type, chemistry and temperature.^7,8^

Although environmental factors play an important role in shaping the distribution of schistosomiasis, transmission is also dependent on human behaviours. The *S. mansoni* life cycle is maintained by faecal excretion of schistosome eggs into freshwater by infected humans, once in the water eggs hatch into miracidia which infect *Biomphalaria* snails then develop into cercariae capable of infecting humans exposed to infested water.^9^ Important risk factors for human infection therefore broadly relate to the frequency and type of contact with infested water sources as well as community-level defecation practices.

In Madagascar, rice farming has been consistently associated with schistosomiasis infection, likely due to occupational water exposure.^10–12^ Rice cultivation is the primary occupation of Madagascar’s agricultural workforce, which account for 74% of the population.^13^ Within rice farming communities, the behavioural and environmental factors associated with infection and intensity of infection, however, remain poorly characterised. Understanding these risk factors is essential to inform targeted control strategies, particularly in Madagascar, where control campaigns through preventative chemotherapy (PC) have proved less effective than elsewhere in sub-Saharan Africa.^7,14^

PC campaigns may have limited effect in Madagascar for multiple reasons, including inadequate resources, widespread poverty and a dispersed rural population with limited transport infrastructure and healthcare access.^7^ The high levels of occupational exposure from rice farming in high burden settings pose a further challenge, as rapid post-treatment re-infection undermines the sustained efficacy of PC.

Surveillance data informing PC planning in Madagascar also remain sparse, with prevalence estimates derived from district-level sentinel surveys that may obscure substantial within-district heterogeneity, risking under-treatment of high-burden communities. Existing prevalence studies have concentrated on the central and northern regions and few include infection intensity.^10,12,15,16^ To our knowledge, no published studies have estimated schistosomiasis prevalence or infection intensity in the South-east, where many districts are classified as low endemicity (Figure 1) yet rice farming and environmental suitability for *S. mansoni* transmission remain high.^6,17^

**Figure 1.**
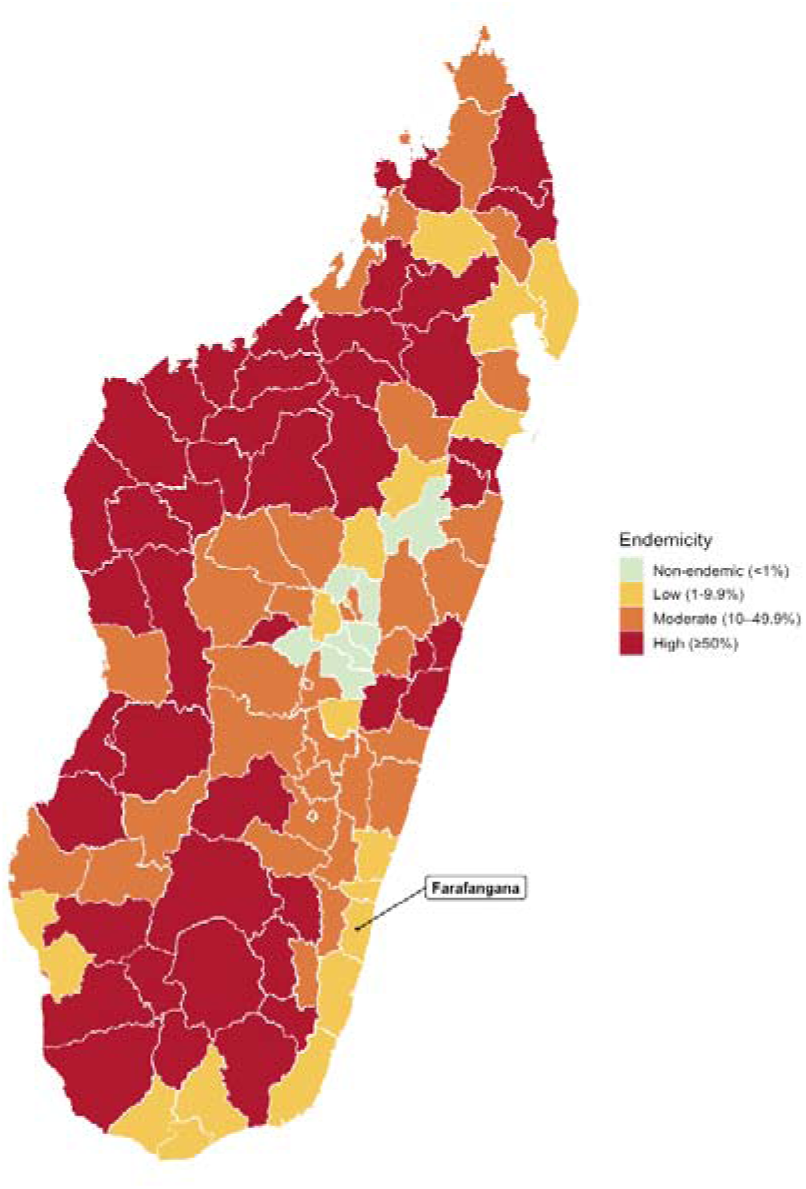
Map of schistosomiasis estimated prevalence in 2024. Data source: WHO African Region: Expanded Special Projected for Elimination of Neglected Tropical Diseases.^17^

The aim of this study was to estimate schistosomiasis prevalence and identify associated behavioural, environmental and WASH-related risk factors in rice farming communities in South-eastern Madagascar.

## Methods

### Study Setting

The Manombo Special Reserve, located in Farafangana District, South-eastern Madagascar, covers approximately 6,633 hectares of protected lowland rainforest and has a population of over 15,000 people living in 31 villages in and around the reserve. Rice farming is the predominant occupation, primarily practiced during the rainy season between November and April. The region has poor road infrastructure constraining programmatic and research activities.

Since 2017, according to the WHO’s Expanded Special Project for Elimination of Neglected Tropical Diseases, the district has had low schistosomiasis endemicity (1-9.9%) with the last PC campaign administered in 2022.^17^ However, a 2023 exploratory unpublished survey conducted by the non-governmental organisation (NGO) Health In Harmony (HIH) using point-of-care circulating cathodic antigen testing, conducted in 2023, found much higher prevalence and schistosomiasis to be endemic in 18 of the 31 villages.

### Study Design

This cross-sectional exploratory study was conducted between 07 May and 31 July 2025 in communities living around the Manombo Special Reserve. Thirteen villages were purposively selected based on relatively high prevalence and sample size within the previous HIH survey. Prior to data collection, the research team visited each of the selected villages to meet with community leaders, explain the study objectives and seek permission to proceed. All village leaders agreed to participate.

### Recruitment

Within selected villages, 12 households were randomly selected for inclusion using HIH-FAMI’s household registry, filtering for households containing at least one adult aged 16 years or older. Within each household, up to two children aged 6 to 15 years and one adult rice farmer aged 16 years or over were included. Where a household contained more than two eligible children or more than one eligible rice farmer, participants were selected by lowest numerical day of birth. Households with neither eligible children nor rice farmers present were excluded. Written informed consent was obtained from all adult participants and from the parents or guardians of participating children, in addition to child assent.

### Data Collection

A structured questionnaire was administered by trained enumerators to adult participants or to child participants’ guardians, recording demographic information, WASH access and practices, geographical locations of water sources and recognised behavioural risk factors for schistosomiasis. Responses were recorded through CommCare (Dimagi, USA), an electronic health record system.

Participants were provided with pre-labelled plastic collection pots and verbal instructions. Caregivers assisted children with sampling. Unless the stool sample could be provided on the day, enumerators returned the following day to collect the samples within 14-16 hours. Stool samples were transported on ice to the field laboratory in Farafangana and stored in the fridge for analysis within 24 hours.

### Laboratory Analysis

Trained laboratory staff prepared three thick-smear Kato-Katz slides from each stool sample, which were read by microscopy.^18^ The number of *S. mansoni* eggs were recorded across all three slides. Soil-transmitted helminth (STH) eggs were also recorded, but results are not presented in this publication. A positive result was defined as detection of one or more eggs on any slide. Infection intensity was calculated as eggs per gram (EPG) of stool by multiplying the arithmetic mean egg counts across the three Kato-Katz slides by a factor of 24.^19^

All participants testing positive for schistosomiasis were offered treatment with praziquantel under supervision of the study clinician.

### Quality Control

To ensure consistency and accuracy of microscopy readings, the study clinician independently re-read 20% of slides, selected at random, blinded to the original result. Discordant readings were reviewed and resolved by consensus. Laboratory staff participated in microscopy training prior to data collection, and slide preparation and reading procedures followed a standardised operating protocol throughout the study.

### Data Analysis

Exposure variables collected through the study questionnaire were selected for inclusion in the analysis based on a prespecified conceptual framework (Appendix 1). Household-level exposures collected from an adult in the household were applied to all children in that household. Clinical and survey data were matched using stool IDs. Records were excluded from analysis if matching could not be conducted or if there were duplicate stool IDs listed.

Since S. *mansoni* can be transmitted through contact with surface water containing infective cercariae and surface water bodies within a catchment are hydrologically connected,^9^ water catchment area boundaries were utilised to delineate potential boundaries of surface water exposure within the study area, an approach informed by prior watershed-based work in this area.^20^ Water catchment area boundaries were obtained from the HydroBASINS Africa Level 07 (v1c) dataset.^21^ Household exposure to each catchment was assigned by spatially linking reported water source GPS coordinates to the corresponding water catchment area. Participants were classified as exposed to a given catchment area if any member of their household used a water source located within it.

Participants were considered infected if any S. *mansoni* eggs were identified in any of the three Kato-Katz slides. Prevalence was estimated overall and by village, participant characteristics, risk factors, water source and water catchment area exposure. Unadjusted relative risks with 95% Wald confidence intervals were estimated for each exposure variable using Poisson regression with robust sandwich standard errors. Infection intensity among Kato-Katz positive participants was summarised as median EPG of stool with interquartile range; differences across exposure groups were assessed using Wilcoxon rank-sum or Kruskal-Wallis tests as appropriate, given the skewed distribution of egg count data.

Multivariable mixed-effects logistic regression was fitted with pre-specified covariates and village as a random intercept to account for clustering. A parallel mixed-effects linear regression model fitted with the same covariate structure and random intercept was used to model log(EPG+1) among Kato-Katz-positive participants. Participants reporting water sources across more than one catchment area were excluded from multivariable analyses. Data were analysed using R version 4.5.2.

## Results

A total of 294 participants from 13 villages were surveyed between May-July 2025, 248 stool samples were collected and analysed by Kato-Katz, and 227 paired survey and stool samples are included in the analysis. Twenty-one stool samples (8.5%) had labelling issues and were excluded from the analysis: 8 had duplicate IDs assigned to different participants; 13 could not be linked to a survey record. Across all 21 excluded samples, 61.9% (13/21) had a positive result for infection.

Participant characteristics are summarised in Table 1. Of the participants included in the analysis, 39.6% (90/227) were male, the median age was 13 years (IQR 9-36) and 58.6% (133/227) were children (<16). Only 34.0% (32/94) of adults were literate with 48.9% (46/94) having never received formal education. Surface water contact was almost ubiquitous with 97.3% (221/227) of participants reporting exposure within the preceding 4 weeks to the study. Among these, bathing was the most common reason for exposure, reported by 97.7% (216/221), followed by crossing a water body 57.9% (128/221) and swimming 41.6% (92/221). Only 5.7% (13/227) of participants had been treated with praziquantel (PZQ) in the six months prior to the survey.

**Table 1.** Prevalence and intensity of S. mansoni infection by demographic, behavioural, and water source exposure characteristics. Unadjusted relative risk with 95% Wald confidence intervals estimated using Poisson regression with robust sandwich standard errors. Infection intensity presented as median EPG (IQR) among positives; intensity p-values from Wilcoxon rank-sum or Kruskal-Wallis tests as appropriate.

|  | n | n Positive | Prevalence | Relative Risk (95% CI) | p value (RR) | Median Eggs per Gram in Positive Samples (IQR) | p value (EPG) |
| --- | --- | --- | --- | --- | --- | --- | --- |
| Socio-Demographics |  |  |  |  |  |  |  |
| Age |  |  |  |  |  |  |  |
| 6-10 | 81 | 44 | 54.30% | ref | ref | 96.0 (29.4–241.8) | 0.963 |
| 11-15 | 49 | 30 | 61.20% | 1.13 (0.84–1.52) | 0.433 | 92.4 (40.8–354.0) |  |
| 16-20 | 5 | 3 | 60.00% | 1.1 (0.53–2.32) | 0.793 | 120.0 (63.6–176.4) |  |
| 21-30 | 16 | 13 | 81.20% | 1.5 (1.1–2.04) | 0.011 | 88.8 (55.2–247.2) |  |
| 31-40 | 32 | 14 | 43.80% | 0.81 (0.52–1.25) | 0.336 | 108.0 (59.4–330.0) |  |
| 41-50 | 23 | 11 | 47.80% | 0.88 (0.55–1.41) | 0.596 | 160.8 (60.0–260.4) |  |
| 51+ | 21 | 6 | 28.60% | 0.53 (0.26–1.06) | 0.074 | 60.0 (35.4–216.0) |  |
| Child | 133 | 75 | 56.40% | ref | ref | 96.0 (36.0–284.4) | 0.623 |
| Adult (16+) | 94 | 46 | 48.90% | 0.87 (0.67–1.12) | 0.276 | 96.0 (49.8–282.0) |  |
| Gender |  |  |  |  |  |  |  |
| Male | 90 | 49 | 54.40% | ref | ref | 160.8 (72.0–376.8) | 0.011 |
| Female | 137 | 72 | 52.60% | 0.97 (0.75–1.24) | 0.779 | 72.0 (31.2–220.2) |  |
| Literate (Adult Only) |  |  |  |  |  |  |  |
| No | 58 | 25 | 43.10% | ref | ref | 72.0 (48.0–288.0) | 0.498 |
| Yes | 32 | 18 | 56.20% | 1.3 (0.85–2) | 0.220 | 111.6 (66.6–259.8) |  |
| Education (Adult Only) |  |  |  |  |  |  |  |
| None | 46 | 19 | 41.30% | ref | ref | 64.8 (48.0–224.4) | 0.340 |
| Primary | 33 | 18 | 54.50% | 1.32 (0.83–2.1) | 0.241 | 132.0 (73.8–366.0) |  |
| Secondary | 7 | 3 | 42.90% | 1.04 (0.41–2.61) | 0.937 | 64.8 (48.0–92.4) |  |
| Higher | 4 | 3 | 75.00% | 1.82 (0.94–3.52) | 0.078 | 247.2 (132.0–255.6) |  |
| PZQ Treatment in prior 6 months |  |  |  |  |  |  |  |
| Not treated | 214 | 112 | 52.30% | ref | ref | 103.2 (40.8–288.0) | 0.707 |
| Treated | 13 | 9 | 69.20% | 1.32 (0.9–1.94) | 0.154 | 72.0 (55.2–88.8) |  |
| Contact with water |  |  |  |  |  |  |  |
| Any Water Contact |  |  |  |  |  |  |  |
| No | 6 | 5 | 83.30% | ref | ref | 103.2 (31.2–151.2) | 0.549 |
| Yes | 221 | 116 | 52.50% | 0.63 (0.43–0.92) | 0.017 | 96.0 (48.0–289.8) |  |
| Fishing |  |  |  |  |  |  |  |
| No | 167 | 88 | 52.70% | ref | ref | 96.0 (48.0–268.2) | 0.644 |
| Yes | 54 | 28 | 51.90% | 0.98 (0.73–1.32) | 0.915 | 92.4 (43.8–391.8) |  |
| Bathing |  |  |  |  |  |  |  |
| No | 5 | 1 | 20.00% | ref | ref | 1152.0 (1152.0–1152.0) | 0.113 |
| Yes | 216 | 115 | 53.20% | 2.66 (0.46–15.43) | 0.275 | 96.0 (48.0–288.0) |  |
| Crossing Water Body |  |  |  |  |  |  |  |
| No | 93 | 55 | 59.10% | ref | ref | 79.2 (39.6–264.0) | 0.225 |
| Yes | 128 | 61 | 47.70% | 0.81 (0.63–1.03) | 0.088 | 127.2 (48.0–312.0) |  |
| Laundry or Washing Dishes |  |  |  |  |  |  |  |
| No | 51 | 27 | 52.90% | ref | ref | 216.0 (68.4–399.6) | 0.055 |
| Yes | 41 | 17 | 41.50% | 0.78 (0.5–1.22) | 0.283 | 64.8 (48.0–96.0) |  |
| Rice Farming |  |  |  |  |  |  |  |
| No | 183 | 100 | 54.60% | ref | ref | 84.0 (46.2–268.2) | 0.085 |
| Yes | 38 | 16 | 42.10% | 0.77 (0.52–1.14) | 0.196 | 216.0 (84.6–522.6) |  |
| Swimming |  |  |  |  |  |  |  |
| No | 129 | 72 | 55.80% | ref | ref | 79.2 (48.0–282.6) | 0.442 |
| Yes | 92 | 44 | 47.80% | 0.86 (0.66–1.11) | 0.250 | 115.2 (38.4–342.0) |  |
| WASH |  |  |  |  |  |  |  |
| Open Defecation (Adults only) |  |  |  |  |  |  |  |
| Never | 21 | 11 | 52.40% | ref | ref | 64.8 (39.6–196.8) | 0.342 |
| Sometimes | 62 | 28 | 45.20% | 0.86 (0.53–1.41) | 0.554 | 99.6 (55.2–251.4) |  |
| Always | 11 | 7 | 63.60% | 1.21 (0.66–2.22) | 0.528 | 384.0 (63.6–456.0) |  |
| Latrine Accessibility |  |  |  |  |  |  |  |
| Accessible | 192 | 101 | 52.60% | ref | ref | 88.8 (40.8–247.2) | 0.090 |
| Inaccessible | 35 | 20 | 57.10% | 1.09 (0.79–1.49) | 0.609 | 240.0 (53.4–515.4) |  |
| Drinking Water Source |  |  |  |  |  |  |  |
| Improved | 30 | 12 | 40.00% | ref | ref | 84.0 (49.2–311.4) | 0.938 |
| Unimproved | 197 | 109 | 55.30% | 1.38 (0.88–2.18) | 0.163 | 96.0 (48.0–288.0) |  |
| Water Catchment |  |  |  |  |  |  |  |
| Household Water Source Location |  |  |  |  |  |  |  |
| Catchment 1 - Not exposed | 140 | 87 | 62.10% | ref | ref | 127.2 (48.0–327.6) | 0.007 |
| Catchment 1 - Exposed | 87 | 34 | 39.10% | 0.63 (0.47–0.84) | 0.002 | 68.4 (24.0–119.4) |  |
| Catchment 2 - Not exposed | 104 | 55 | 52.90% | ref | ref | 88.8 (48.0–156.0) | 0.223 |
| Catchment 2 - Exposed | 123 | 66 | 53.70% | 1.01 (0.79–1.3) | 0.907 | 144.0 (31.2–366.6) |  |
| Catchment 3 - Not exposed | 188 | 92 | 48.90% | ref | ref | 96.0 (31.2–299.4) | 0.768 |
| Catchment 3 - Exposed | 39 | 29 | 74.40% | 1.52 (1.2–1.92) | <0.001 | 96.0 (55.2–232.8) |  |

Open defecation in the seven days prior to the survey was reported as “always” by 11.7% (11/94) and “sometimes” by 65.9% (62/94) of adult participants. Of these, 80.8% (59/73) reported practicing open defecation in the forest, 27.4% (20/73) on the ground in the village and 5.5% (4/73) in or near agricultural plots. Most participants had access to unlined pit latrines (192/227, 84.6%) and used unimproved water sources as a primary drinking water source (197/227, 86.7%). Having access to a tap in the household or compound was uncommon (9/227, 4.0%). There was no significant variation in open defecation rates at a village level.

All 13 villages and 31 water sources were located across three water catchment areas. Participants mainly used water sources in the same catchment area as their village. However, 4.4% (10/227) used water sources across two catchment areas.

Among the included 227 participants, the prevalence of *S. mansoni* infection was 53.3% (121/227, 95% CI 46.8–59.8%). Among infected participants, median infection intensity was 96 (IQR 48 – 288) EPG. Prevalence was similar between males and females, although males had a higher infection intensity than females. In univariable analyses, there was no strong evidence of association between infection status or infection intensity and PZQ treatment in the preceding 6 months, demographic, behavioural or WASH-related covariates.

Prevalence and intensity of *S. mansoni* infection varied considerably by village with spatial clustering observed among both villages and water sources (Figure 2). Participants reporting water contact in catchments 2 and 3 were significantly more likely to be infected than those in catchment 1. These associations persisted in multivariable analyses (n=203; participants with water sources spanning more than one catchment or with missing exposure or covariate data were excluded) adjusting for age, sex, and PZQ treatment within the preceding 6 months, village level open defecation rates and village random effects (Figure 3). Infection intensity was also higher among infected individuals in catchments 2 and 3, however this difference was not statistically significant.

**Figure 2.**
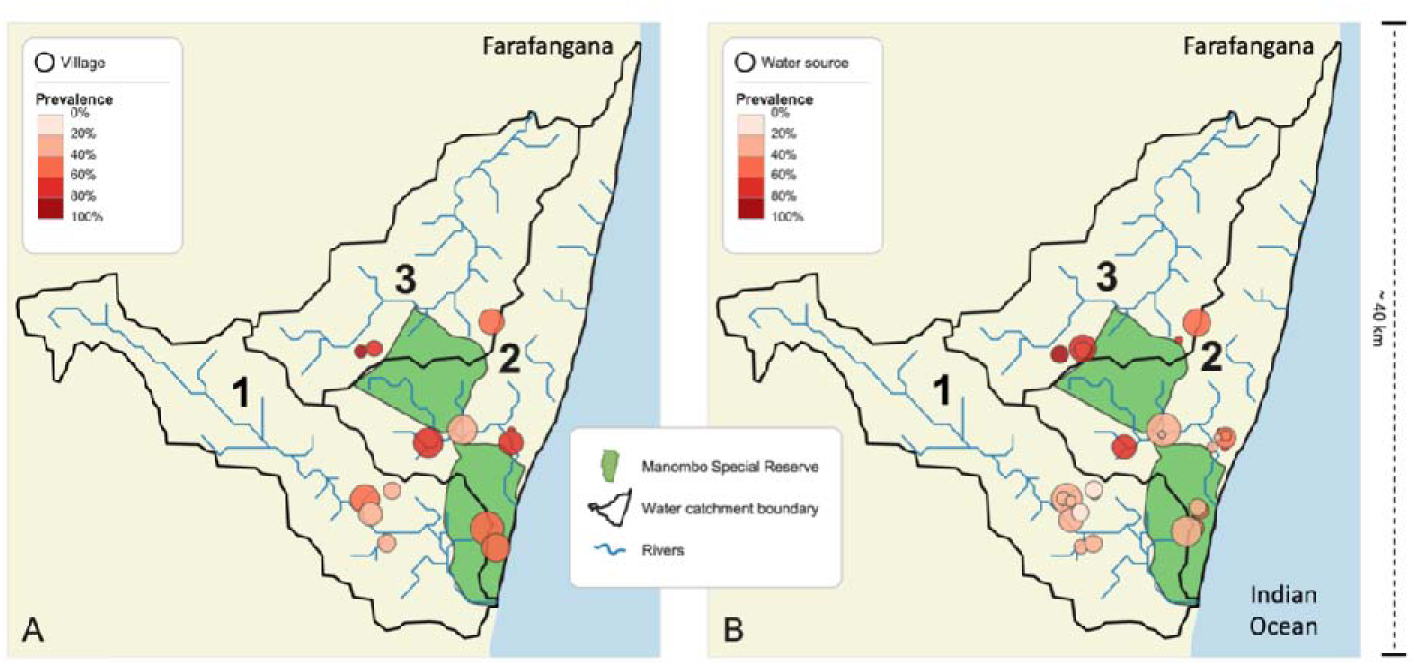
A - Schistosomiasis prevalence among residents of 13 villages, B - Schistosomiasis prevalence among users of 31 water sources. Labels 1-3 denote water catchment areas. Circle size is proportional to the number of residents (A) or water source users (B). Darker red indicates higher prevalence amongst residents (A) or water source users (B).

**Figure 3.**
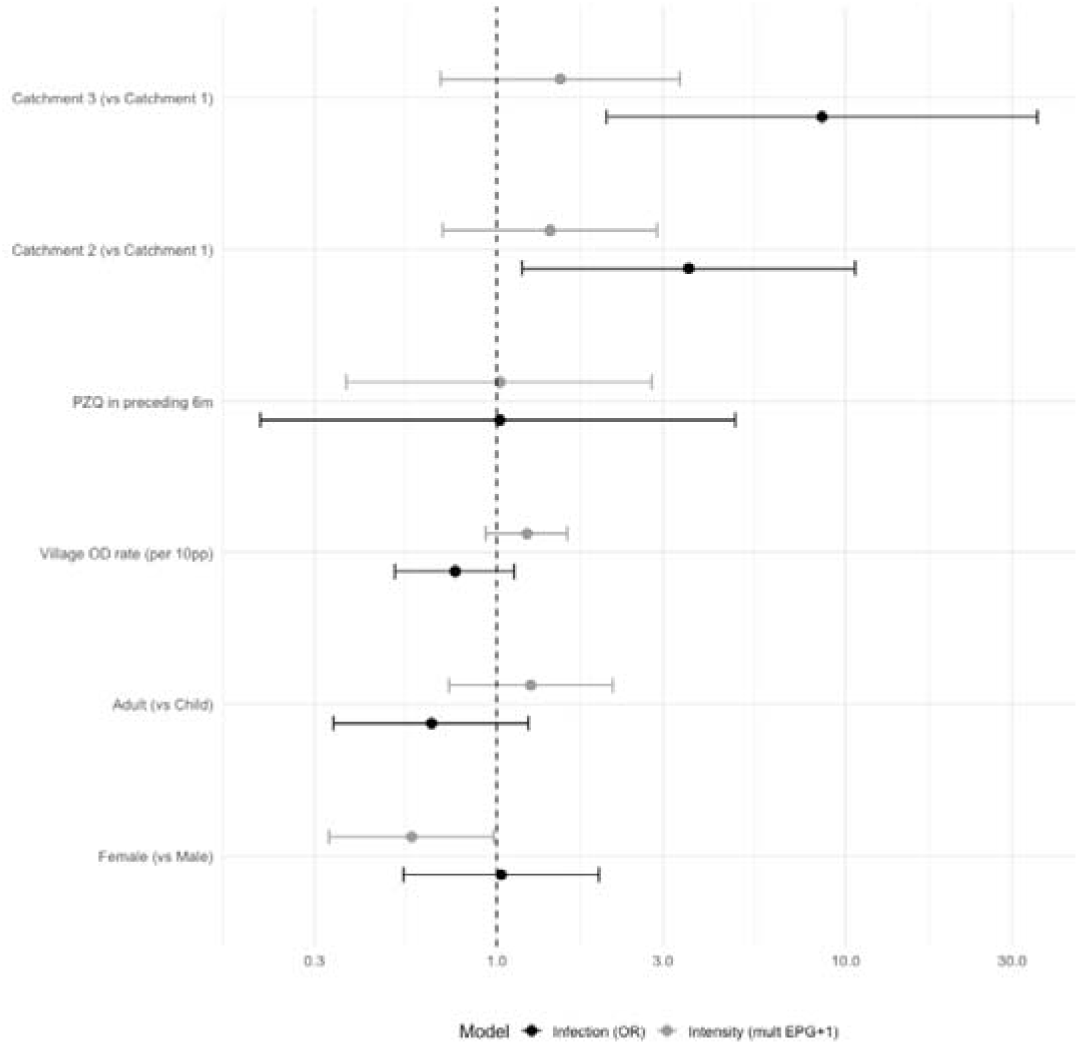
Forest plot of multivariable model estimates, odds ratio of infection and intensity ratio (EPG+1 multiplicity) with 95% CI. OD = Open Defecation. Catchment 1 used as water source location reference.

## Discussion

To our knowledge, this is the first study assessing *S. mansoni* infection and intensity in rice farming communities in South-eastern Madagascar. Overall, prevalence was consistent with high endemicity,^22^ but varied substantially between villages. Spatial clustering of infection prevalence was observed and associated with water catchment area boundaries. Surface water contact was nearly universal among participants and there was no strong evidence of associations with demographic characteristics, water contact behaviour, prior PZQ treatment, village level open-defecation rates or WASH access indicators. Collectively, these findings suggest that transmission risk in this setting is associated with environmental conditions rather than individual-level behavioural exposures or household WASH access.

### Water catchment areas as a determinant of infection

The association between infection risk and water catchment area is supported by a plausible biological mechanism. Water catchments areas are discrete hydrological units within which precipitation collects and drains through a connected stream network to a common outlet. A catchment area can determine the characteristics of the water bodies including flow velocity, depth, water chemistry, biodiversity and flood frequency, duration, extent, and intensity.^23–27^

The catchments in the study area are similar in size (ranging from 157 to 191 km²) but differ in stream-network structure. Catchments 1 and 2 each contain perennial fifth-order streams (the Karimbelo and Tokoandra Rivers, respectively), whereas the water sources documented in catchment 3 are limited to intermittent, low-order tributaries that flow seasonally. Across the subset of sites sampled in a prior dry-season survey of this study area, pH (means ranging from 5.4 to 6.1), specific conductance (mean < 70 µS/cm), temperature, and major ion concentrations were broadly similar among catchments.^20^ The few outliers reflect site-specific conditions, with higher salt concentrations at the lowest reach of the Karimbelo River where it approaches the Indian Ocean and elevated manganese at two sources in catchment 1, rather than catchment-scale differences. The catchment effect on infection prevalence is therefore unlikely to be explained by differences in water chemistry. A more plausible explanation lies in the physical character of the water bodies.

*Biomphalaria* snails are more abundant in shallow water.^25^ Their habitat suitability is also associated with marshy, vegetated conditions and shifts seasonally with flooding, conditions typical of the seasonal low-order tributaries and adjacent rice paddies in catchment 3 which exhibited the highest prevalence of infection.^27^ The perennial higher-order rivers in catchments 1 and 2 are less consistent with this habitat profile, however this mechanism does not explain the prevalence difference between these two catchments. Further analysis of water velocity, channel depth, and snail abundance would be needed to support a mechanistic link between catchment and infection prevalence and represent a priority for future work.

### Heterogeneity of prevalence

Infection prevalence varied markedly across the 13 villages located within a few kilometres of each other, ranging from 15% to 100%, though sample sizes per village were small. Surveillance systems relying on a small number of sentinel sites may not adequately capture heterogeneity existing at such a fine geographical scale, potentially explaining the substantial discrepancy between the 56.4% prevalence among school-aged children (SAC) observed in this cohort and the current low endemicity (1-9.9%) designation for the study district based on national sentinel surveillance estimates.^17^

Standard sentinel surveillance programmes derive prevalence estimates from school-based sampling of SAC. Within the study population, 48.9% of adult participants reported no formal education and 34.0% were literate, far below the national average of 75%.^3^ This is likely indicative of relatively low school attendance in this community. Where school enrolment is consistently low, school-based surveillance may systematically exclude those communities with greatest exposure such as those engaged in rice farming. This represents a potential structural inequity whereby the populations at highest transmission risk are least likely to be captured by standard prevalence estimation methodology and therefore least likely to receive treatment, a risk acknowledged by the WHO.^28^

### Behavioural and WASH related factors

Commonly cited risk factors for schistosomiasis were not significantly associated with infection or intensity in this cohort. This is likely influenced by uniformly high levels of surface water contact, with 97.7% of participants reporting regular use of surface water for bathing, which limited variability in behavioural exposure and reduced the ability to detect associations with individual-level risk factors. Where water contact is ubiquitous, variation in infection risk may be more strongly shaped by features of the transmission environment, such as water source characteristics, than by individual behaviour or demographic factors. However, water contact was only recorded as a binary variable, precluding assessment of exposure frequency, duration, or intensity. This limitation, combined with a small sample size, may have obscured dose-dependent or more subtle associations.

Our results found that while there are no differences in infection rates, infection intensity was higher in men than women, possibly due to increased occupational exposure during rice farming, fishing, or irrigation activities, which typically involve more prolonged and whole-body water contact than the domestic use more commonly undertaken by women, consistent with findings from other studies in Madagascar.^10^

### Implications for surveillance

Our findings of high prevalence heterogeneity and spatial clustering associated with water catchment areas provide an avenue warranting further investigation. When administrative boundaries are used to define sentinel survey areas, these may encompass hydrologically distinct zones with substantially different transmission risk, without diverse sampling across water catchment areas. Water catchment areas, which align more closely with the ecological determinants of transmission, could represent more representative sampling units within districts and highlight specific areas for targeted intervention. However, further research is needed to evaluate the practicality and legitimacy of this approach.

### Implications for control

Given that there are very high levels of surface water exposure both through occupation and activities of daily living, behaviour change interventions aimed at reducing water contact may not be feasible or effective. While most households had access to latrines, open defecation was still common, mostly occurring outside the confines of the villages. The reasons for this are likely multifactorial, but one possible explanation is that open defecation occurs mainly while individuals are farming, rather than at home. The implications of open defecation for schistosomiasis transmission may also differ according to the setting in which it occurs and the relative contribution of open defecation in agricultural areas compared with household environments remains unclear. Understanding the barriers to latrine use in this context, whether structural, cultural or related to maintenance and acceptability, is a prerequisite for designing effective interventions and may represent a more tractable avenue for transmission control than reducing water contact.

Snail control is another alternative modality of interruption of transmission. A recent malacological survey in the region failed to identify *Biomphalaria* species at any sampled site; our results suggest that water bodies in catchments 2 and 3 deserve closer malacological investigation and possible location for vector control strategies.^29^

Finally, PC may be an underutilised strategy for this community. Recent treatment was very rare amongst participants and was linked to previous research surveys rather than routine healthcare diagnosis and treatment. Between 2014 and 2024 there have been three documented PC campaigns in Farafangana district.^17^ This community was, however, not reached. In addition, the high burden and regular water contact may limit effectiveness through rapid reinfection.

### Limitations

This exploratory study was conducted in villages selected based on expected high *S. mansoni* prevalence, therefore reducing generalisability to the wider Farafangana district. The relatively small sample size further limits the precision of prevalence estimates and the power of the multivariable analysis, particularly for the smaller exposure groups such as catchment 3. Stool samples were collected on a single day and read by a single analyst without formal quality control, which may have reduced sensitivity, particularly for light infections, and may have resulted in an underestimate of true prevalence.

This was a cross-sectional study and therefore causal inference regarding water catchment exposure and S. *mansoni* infection is not possible. The observed associations may reflect unmeasured confounding by factors correlated with catchment geography, such as proximity to water bodies, irrigation infrastructure or rice farming intensity. Furthermore, as only prevalent infection was measured, we cannot exclude the possibility that current infection status reflects historical exposures not captured by the study, particularly given the long lifespan of adult schistosomes.

## Conclusions

*S. mansoni* prevalence in this rice farming community in a rural area of South-eastern Madagascar substantially exceeded current district-level estimates, with marked variation between neighbouring villages. This suggests that transmission intensity can vary considerably over very short distances in this setting, likely reflecting fine-scale ecological heterogeneity that is not captured by routine surveillance frameworks. Differing exposure to water catchment area was the most strongly associated environmental factor with infection, potentially consistent with known *Biomphalaria* habitat preferences. These findings suggest that hydrological factors may play an important role in shaping transmission heterogeneity in Madagascar, warranting further malacological and epidemiological investigation.

## Data Availability

Data produced in the present study are available upon reasonable request to the authors.

## Declarations

### Ethics approval and consent to participate

Ethical approval was obtained from the London School of Hygiene and Tropical Medicine Ethics Ref: 31176 and Comité Malgache d’Éthique pour les Sciences et les Technologies: Project CcHy-St.

### Competing Interests

The authors declare that they have no competing interests

### Funding

This project was supported by an unrestricted donation from Reckitt plc, DONAT16113, to the London School of Hygiene and Tropical Medicine to support research and learning on hygiene and health. The funder had no input on the study design, methods, or decision to publish.

### Authors Contributions

James Bevan: Data curation; Formal analysis; Visualisation; Writing – original draft; Writing – review & editing. Nina Finley: Conceptualisation; Methodology; Investigation; Writing – review & editing. Herizo Randrianandrasana: Data curation; Investigation; Resources. Tanguy Felistino Razafiarimihoby: Investigation. Andry Tsirimanana: Investigation; Resources. Elinambinina Rajaonarifara: Data curation; Resources. Ryan Parks: Writing – review & editing; Data curation; Visualisation. Lena Reiter: Data curation. Tahinamandranto Rasamoelina: Conceptualisation. Sakib Burza: Conceptualisation; Writing – review & editing. Laura Braun: Conceptualisation; Methodology; Investigation; Supervision; Writing – review & editing.

## Acknowledgements

We extend our gratitude to the participants and communities for their willingness to contribute to this research. We thank our field enumerators, Rakotoson Heridahy Felistin and Baoarivony Anny Vannessa, for their dedication throughout data collection. The support of Health In Harmony in facilitating access to study communities in Farafangana District is gratefully acknowledged.

James Bevan, NIHR Academic Clinical Fellow, ACF-2024-20-004, is funded by Health Education England (HEE) / NIHR for this research project. The views expressed in this publication are those of the author(s) and not necessarily those of the NIHR, NHS or the UK Department of Health and Social Care."

**Appendix 1:**
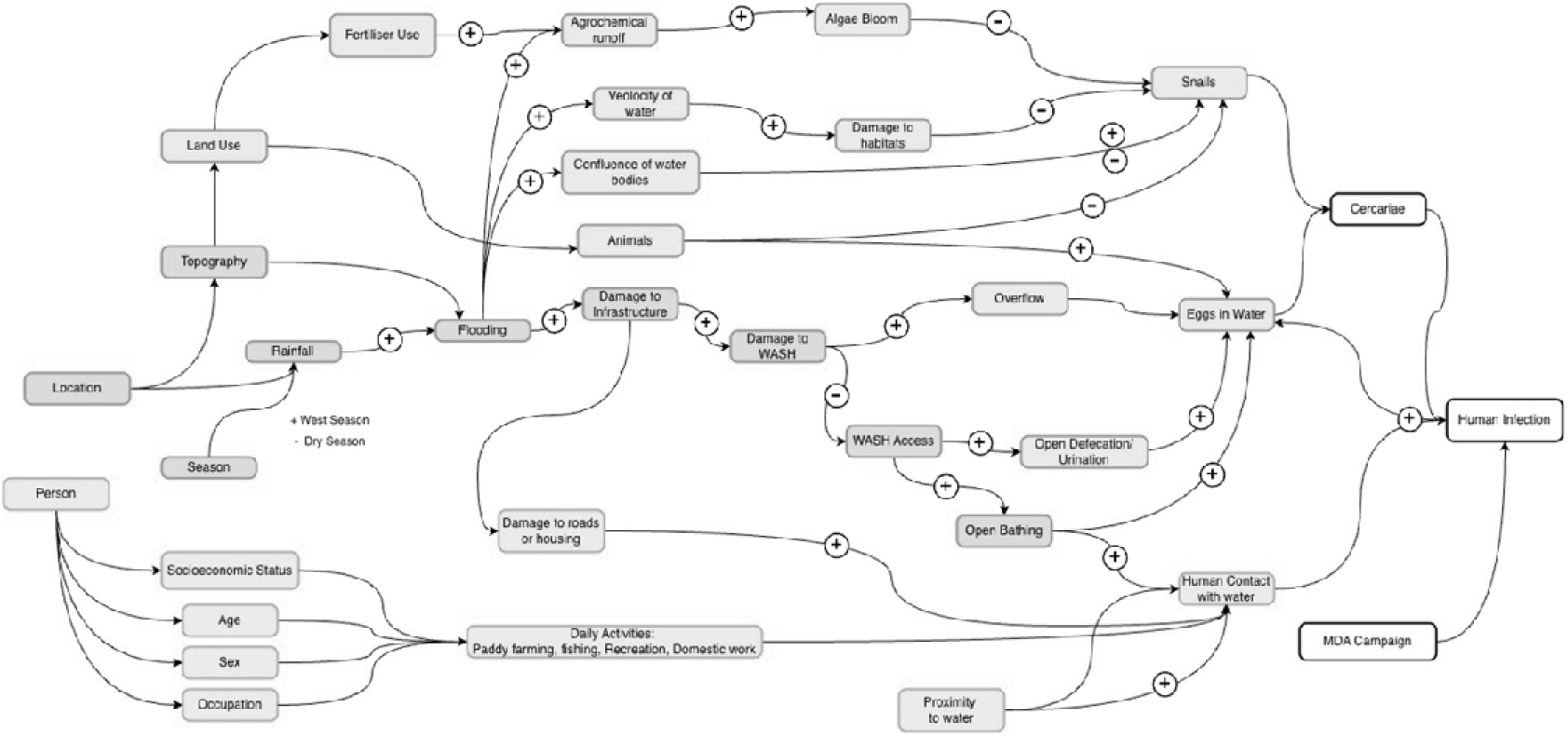
Conceptual framework of demographic, behavioural, environmental and WASH access related exposures associated with schistosomiasis infection.

**Appendix 2.** Multivariable model estimates, odds ratio of infection (logistic regression) and intensity ratio (EPG+1 multiplicity)

| section | Exposure | OR | P value | int_mult_CI | P value |
| --- | --- | --- | --- | --- | --- |
| Water source catchment area | Catchment 1 | 1.00 | NA | 1.00 | NA |
|  | Catchment 2 | 3.12 (1.28–7.59) | 0.012 | 1.43 (0.7–2.95) | 0.33 |
|  | Catchment 3 | 4.41 (1.63–11.94) | 0.003 | 1.51 (0.69–3.32) | 0.301 |
| Gender (ref: Male) | Female | 0.98 (0.57–1.68) | 0.943 | 0.56 (0.32–0.96) | 0.037 |
| Child/Adult (ref: Child) | Adult | 0.86 (0.5–1.49) | 0.587 | 1.26 (0.74–2.18) | 0.396 |
| Treatment | Treated in preceding 6m | 1.81 (0.54–6.07) | 0.339 | 1.02 (0.37–2.77) | 0.975 |
| WASH | Village open defecation rate (per | 0.89 (0.66–1.21) | 0.457 | 1.2 (0.92–1.57) | 0.188 |
|  | 10pp) |  |  |  |  |

## Notes

### Competing Interest Statement

The authors have declared no competing interest.

### Author Declarations

Ethical approval was obtained from the London School of Hygiene and Tropical Medicine Ethics Ref: 31176 and Comite Malgache d'Ethique pour les Sciences et les Technologies: Project CcHy-St.

